# Effectiveness of Covid-19 Vaccines against the SARS-COV-2-Delta (B.1.617.2) in China-A Real World Study

**DOI:** 10.1101/2022.02.07.22270490

**Authors:** Xinge Ma, Jianfeng Han, Hongxia Li, Chang Liu

## Abstract

**Background:** Severe acute respiratory syndrome coronavirus 2 (SARS-CoV-2) delta (B.1.617.2) variant is highly transmissible and has contributed to a surge in cases globally. This study aimed to explore the potential of vaccines against SARS-CoV-2 delta (B.1.617.2) variant in China.

**Methods:** In this real-world study, all data were extracted from Xi’an Chest Hospital. Confirmed cases infected with Delta VOC with exact date of positive viral testing were included for analysis. Patients meeting the study criteria were divided into unvaccinated and partially vaccinated (one dose), full vaccinated (two doses), and booster vaccination of COVID-19.

**Results:** A total of 455 cases were enrolled in this study. Proportion of severe and critical cases in full vaccinated cases (1.82%) and cases with booster vaccination (1.35%) of COVID-19 were much lower than that of unvaccinated and partially vaccinated cases (8.16%). In addition, cases with booster vaccination (12.78 days) and full vaccinated cases (12.59 days) showed shorter duration of viral shedding than that in unvaccinated and partially vaccinated cases (13.87 days).

**Conclusion:** This is the first real world study indicating that Covid-19 vaccines showed much powerful effectiveness against the SARS-COV-2-Delta (B.1.617.2) in China, including lowing the proportion of severe illness and shorting the virus shedding time.

## Introduction

Severe acute respiratory syndrome coronavirus 2 (SARS-CoV-2) delta (B.1.617.2) variant is highly transmissible and has contributed to a surge in cases globally.^1-3^ The effectiveness of the Chinese vaccines against this variant in Chinese population remains unknown.

## Methods

Since the identification of the first locally transmitted Delta variant case connected to an imported case on December 9, 2021, all patients with positive RT-PCR test of COVID-19 were in three designated hospitals. Confirmed cases infected with Delta VOC with exact date of positive viral testing and decline in the largest COVID-only designated hospital of Xi’an Chest Hospital were included for clinical characteristics and duration of viral shedding analysis among cases with unvaccinated and partially vaccinated (one dose), full vaccinated (two doses), and booster vaccination of COVID-19. This study was approved by the local institutional research and ethics committee (XJTU1AF2022LSK-001).

## Results

All cases during this outbreak in Xi’an were identified as a cluster having the same nucleic acid sequence as the Delta VOC. A total of 455 cases infected with exact date of both positive viral testing and decline in Xi’an Chest Hospital were included in this study, proportion of severe and critical cases in full vaccinated cases (1.82%) and cases with booster vaccination (1.35%) of COVID-19 were much lower than that of unvaccinated and partially vaccinated cases (8.16%) (Figure 1). Cases with booster vaccination (12.78 days) and full vaccinated cases (12.59 days) showed shorter duration of viral shedding than that in unvaccinated and partially vaccinated cases (13.87 days) (Figure 2).

**Figure 1.**
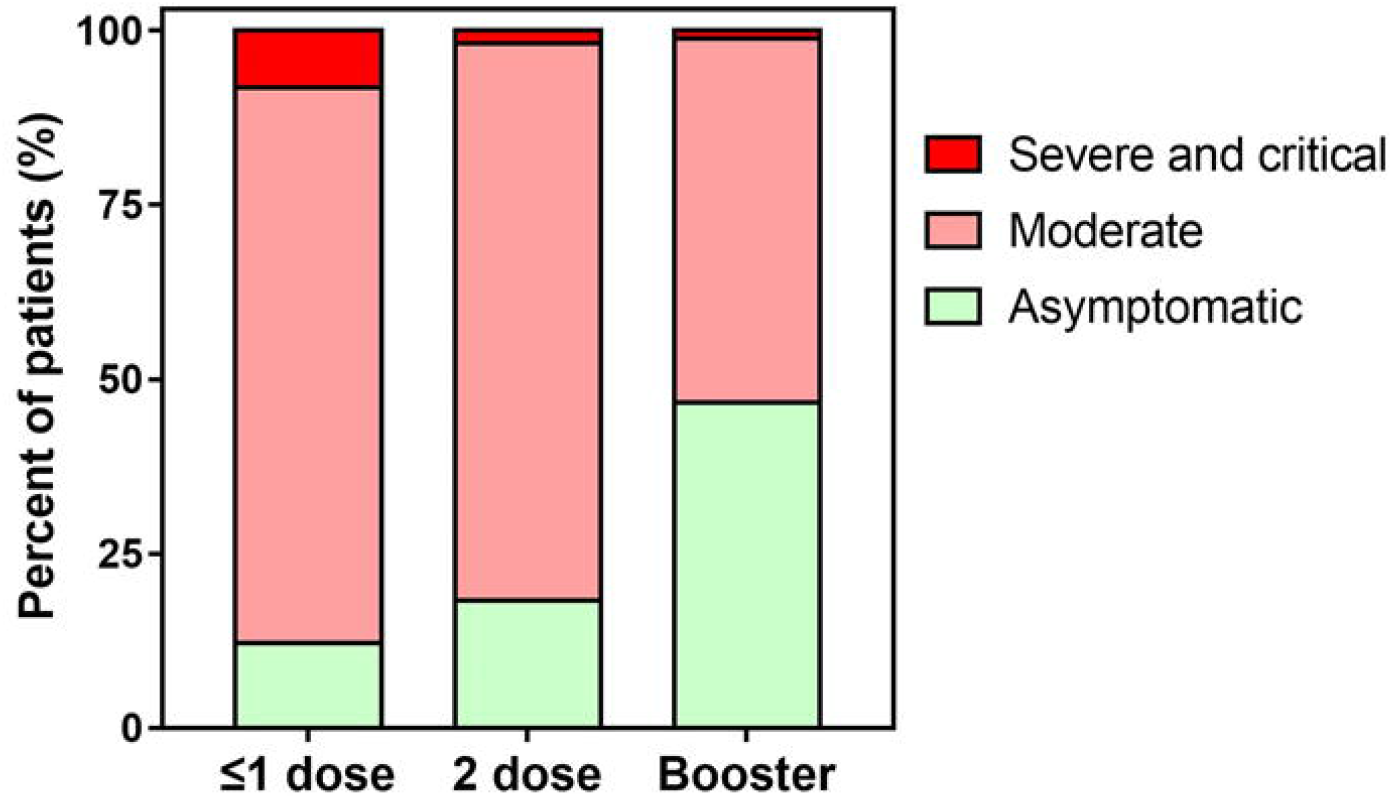
The potential of vaccination in reducing the proportion of severe and critical cases.

**Figure 2.**
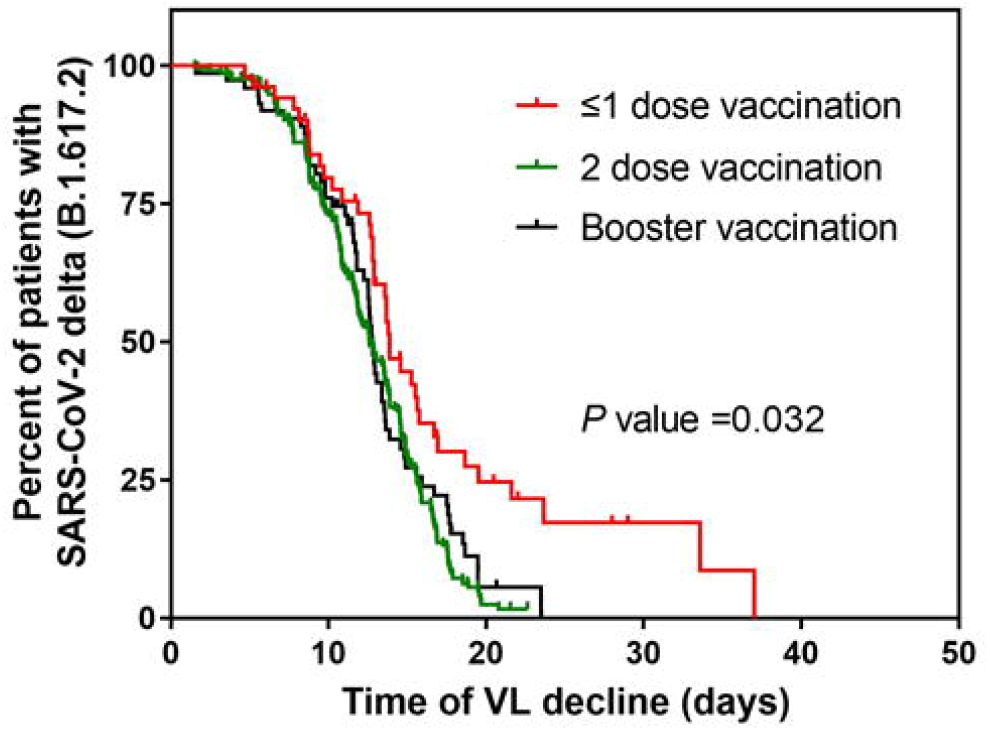
The potential of vaccination in shorting the duration of viral shedding.

## Discussion

To our knowledge, this is the first real-world study which has focused on the effect of vaccination against the delta variant in Chinese population in a local outbreak. Our study showed powerful potential of two doses or booster vaccination in reducing the proportion of severe or critical cases and shorting the duration of viral shedding.

All locally transmitted cases during this local outbreak in Xi’an were connected to an imported case between December 9, 2021 (the first case officially confirmed) and January 20th, 2022. After at least six rounds of citywide viral RNA testing (including a total of 100 million people), all confirmed cases were admitted in the COVID-only designated hospitals and some had the exact date of positive viral testing and decline. Moreover, the COVID-19 vaccination coverage in adult people of Xi’an was 95.51%. These exact data provided us a clear overall picture to better understand the effect of vaccines against the delta variant in Chinese population. First, vaccines showed powerful potential in reducing the proportion of severe or critical cases. Severe or critical COVID-19-recovered cases can develop physical and psychological impairments during follow-up.^4^ Therefore, reducing the proportion of severe or critical cases who require intensive care unit admission is widely considered to one, if not the most important, factor to determine whether the campaigns against COVID-19 can be successful. Second, these findings also suggest the necessity of the original policy decision on dosing schedule especially for those more susceptible to the infection with the Delta VOC. Since COVID-19 outbreak in December 2019, there have been several globally circulating SARS-CoV-2 variant strains. During the study period, a novel variant of SARS-CoV-2, the Omicron variant of concern, is fast becoming the dominant strain globally, including China, and has resulted in a considerable socioeconomic burden. We should all stand together against the COVID-19 pandemic and be carefully identified to take more actions, especially vaccination dosing schedule, before the condition deteriorates.

## Limitations

Some limitations should be noted. The limited numbers of cases are currently insufficient to make a subgroup comparison of the effectiveness of different kinds of vaccines. Second, there are no follow-up data to evaluate the long-term vaccine protection against physical and psychological disorders. The findings are observational and should be interpreted with caution.

## Data Availability

All data produced in the present study are available upon reasonable request to the authors.

